# Medication-related adverse events in patients with cancer and discrepancies in cystatin C- versus creatinine-based eGFR

**DOI:** 10.1101/2023.01.18.23284656

**Authors:** Paul E. Hanna, Qiyu Wang, Ian Strohbehn, Daiana Moreno, Destiny Harden, Tianqi Ouyang, Nurit Katz-Agranov, Harish Seethapathy, Kerry L. Reynolds, Shruti Gupta, David E. Leaf, Meghan E. Sise

**Affiliations:** Division of Nephrology, Department of Medicine, Massachusetts General Hospital, Boston, MA; Division of Hematology and Oncology, Department of Medicine, Massachusetts General Hospital, Boston, MA; Division of Renal Medicine, Brigham and Women’s Hospital, Boston, MA; Adult Survivorship Program, Dana-Farber Cancer Institute, Boston, MA

## Abstract

**Background:** Creatinine-based estimated glomerular filtration rate (eGFR_CRE_) may overestimate kidney function in patients with cancer. Cystatin C-based eGFR (eGFR_CYS_) is an alternative marker of kidney function. We investigated whether patients with an eGFR discrepancy, defined as eGFR_CYS_ >30% lower than the concurrent eGFR_CRE_, had an increased risk of adverse events resulting from renally-cleared medications.

**Patients and Methods:** We conducted a cohort study of adult patients with cancer who had serum creatinine and cystatin C measured on the same day between May 2010 and January 2022 at two academic cancer centers in Boston, MA. The primary outcome was the incidence of each of the following medication-related adverse events: 1) supratherapeutic vancomycin levels (>30μg/mL); 2) trimethoprim-sulfamethoxazole-related hyperkalemia (>5.5mEq/L); 3) baclofen-induced neurotoxicity; and 4) supratherapeutic digoxin levels (>2.0ng/mL).

**Results:** 1988 patients with cancer had simultaneous eGFR_CYS_ and eGFR_CRE_. The mean age was 66 years (SD±14), 965 (49%) were female, and 1555 (78%) were non-Hispanic white. eGFR discrepancy occurred in 579 patients (29%). Patients with eGFR discrepancy were more likely to experience medication-related adverse events compared to those without eGFR discrepancy: vancomycin levels >30μg/mL (24% vs. 10%, p=0.004), trimethoprim-sulfamethoxazole-related hyperkalemia (24% vs. 12%, p=0.013), baclofen-induced neurotoxicity (25% vs. 0%, p=0.13), and supratherapeutic digoxin levels (38% vs. 0%, p=0.03). The adjusted OR for vancomycin levels >30μg/mL was 2.30 (95% CI 1.05 – 5.51, p = 0.047).

**Conclusion:** Among patients with cancer with simultaneous assessment of eGFR_CYS_ and eGFR_CRE_, medication-related adverse events occur more commonly in those with eGFR discrepancy. These findings underscore the importance of accurate assessment of kidney function and appropriate dosing of renally-cleared medications in patients with cancer.

## Introduction

Accurate assessment of estimated glomerular filtration rate (eGFR) is key to dosing renally-cleared medications. While the gold standard method for evaluating kidney function is direct measurement of glomerular filtration rate (mGFR) using inulin or chromium-51 labeled ethylenediamine tetra-acetic acid,^1^ GFR estimation using serum creatinine is the most commonly used method in both clinical practice and research.^2-4^ Creatinine is a byproduct of muscle metabolism that is filtered and secreted by the kidneys. Despite continued improvements of currently available eGFR equations, creatinine-based eGFR remains imprecise and can overestimate kidney function, particularly in patients with sarcopenia.^5, 6^ This can lead to inaccurate dosing of medications that require adjustment based on eGFR, such as commonly used antibiotics, muscle relaxants, anti-epileptic drugs, blood thinners, and antiarrhythmic medications.

Cystatin C is a low molecular weight (13K Dalton) protein produced by all nucleated cells. It is freely filtered by the glomerulus and does not undergo reabsorption or secretion.^7^ Unlike creatinine, cystatin C is not readily affected by age, sex, muscle mass, or diet, and has been increasingly used as an alternative to creatinine to estimate GFR.^2, 5, 8^ A recent, large study in patients with solid tumors demonstrated that using an equation that combines both creatinine and cystatin C is the most accurate way to estimate GFR.^8, 9^

Because cancer is a significant risk factor for sarcopenia,^10^ we hypothesized that having a cystatin C-based eGFR (eGFR_CYS_) that is significantly lower than creatinine-based eGFR (eGFR_CRE_) would be common in patients with cancer. Given that patients with cancer are commonly exposed to numerous medications that require dose adjustment by kidney function, we hypothesized that adverse events related to renally-cleared medications would be higher in patients with a large discrepancy between eGFR_CYS_ versus eGFR_CRE_.

## Methods

### Patient population

Using Mass General Brigham’s centralized data warehouse, the Research Patient Data Registry (RPDR)^11, 12^, we identified adult patients with a pre-existing diagnosis of malignancy who had both serum creatinine and cystatin C measurements on the same day between May 2010 and January 2022. eGFR_CRE_ was calculated using the CKD Epidemiology Collaboration (CKD-EPI) 2021 race-free equation,^5^ while eGFR_CYS_ was calculated using the CKD Epidemiology Collaboration (CKD-EPI) 2012 race-free equation.^13, 14^

### Data collection

The date of the first simultaneous eGFR_CRE_ and eGFR_CYS_ measurement was considered the baseline date. Comorbidities were defined based on diagnosis codes appearing any time prior to the baseline date, and concurrent medication use was defined by active prescription within 1 year prior to the baseline date. Cancer type was determined by the most frequently used cancer-related diagnosis code prior to the baseline. Baseline chronic kidney disease was defined by the 2021 race free CKD-EPI equation that incorporates both serum creatinine and cystatin C ^8^, and chronic kidney disease was staged per Kidney Disease Improving Global Outcomes (KDIGO) guidelines.^15^ Clinical diagnoses of medication adverse events were determined by manual chart review by two investigators; with a third available to resolve disagreement.

### Primary exposure

The primary exposure was eGFR discrepancy, defined as eGFR_CYS_ more than 30% lower than the eGFR_CRE_; the reference group consisted of all other patients and included patients whose eGFR_CYS_ was more than 30% greater than eGFR_CRE_ as this would not place patients at risk factor for adverse medication side effects from renally-cleared medications. The 30% cut-off was chosen because it is commonly used in clinical studies to define the accuracy of eGFR from measured GFR.^9, 16^ We additionally identified a subset of patients with severe eGFR discrepancy, defined as eGFR_CYS_ more than 50% lower than eGFR_CRE_ and eGFR_CYS_ less than 30 mL/min/1.73m^2^.

### Primary outcome: Adverse events related to renally-cleared medications

We examined the risk of selected medication adverse events using detailed chart review. We selected medications (intravenous vancomycin, trimethoprim-sulfamethoxazole, baclofen, and digoxin) that are typically dose-adjusted based on eGFR and whose side effects could be quantified by drug level monitoring, laboratory abnormalities, or identified by chart review. In all cases, we evaluated drug exposures that occurred within 90 days of the baseline date.

The therapeutic range for a vancomycin trough level is 15-20μg/mL and levels greater than 20μg/mL are considered supratherapeutic^17, 18^. We defined severely elevated vancomycin trough levels as those greater than >30 μg/mL and used manual chart review to exclude peak values.^19-21^

Trimethoprim-sulfamethoxazole-related hyperkalemia was defined as a serum potassium level >5.5mEq/L (Common Terminology Criteria for Adverse events [CTCAE v 4.0] grade 2), and severe hyperkalemia was defined as a level >6.0mEq/L (grade 3) within 30 days of starting trimethoprim-sulfamethoxazole. As a sensitivity analysis, we determined the average rise in potassium after initiation of trimethoprim-sulfamethoxazole and the rate of an absolute increase in serum potassium ≥0.5mEq/L from baseline.

Baclofen toxicity was determined by chart review. Baclofen toxicity was defined as altered mental status, myoclonus, seizure, or orthostatic hypotension/dizziness warranting discontinuation of the medication.^22, 23^ An investigator blinded to cystatin C values evaluated all clinical documentation within 90 days of baseline to identify potential cases of baclofen toxicity.

Digoxin toxicity was determined by chart review. An investigator blinded to cystatin C values evaluated all clinical documentation and digoxin levels obtained within 90 days of baseline. Digoxin toxicity was defined as altered mental status, nausea, orthostatic hypotension, or bradycardia attributed to digoxin by the treating team, with a corresponding digoxin trough level above the therapeutic range.^24, 25^

### Secondary outcomes

We evaluated eGFR discrepancy and severe eGFR discrepancy as dependent variables and determined which baseline characteristics and laboratory studies were associated with eGFR discrepancy.

We evaluated the effect of eGFR discrepancy on 30-day mortality. Date of simultaneous eGFR_CRE_ and eGFR_CYS_ served as day 0. Patients lost to follow-up within 30 days were censored at their last visit.

### Statistical Analysis

We reported baseline characteristics using counts and percentages for categorical variables and means and standard deviations (SD) for normally distributed continuous variables, and median and interquartile range for skewed variables. Logistic regression models were used to examine the association between baseline demographics, comorbidities, medications, laboratory studies, and eGFR discrepancy in a univariable model. Serum albumin and hemoglobin were evaluated in clinically relevant categories shown in **Table 1**. Variables were then selected based on clinical plausibility and information criteria (Akaike and Bayesian) to generate the final multivariable model. The Wald Chi-squared test was used to assess the significance of explanatory variables. The final model was adjusted for age, sex, race, eGFR_CRE-CYS_, BMI, smoking, hypertension, coronary artery disease, diabetes, cirrhosis, malnutrition, thyroid disease, proton pump inhibitor use, diuretic use, angiotensin converting enzyme inhibitor or angiotensin receptor blocker use, corticosteroid use, serum albumin, and hemoglobin,

**Table 1.** Patient Characteristics. eGFR discrepancy was defined as eGFR_CYS_ more than 30% lower than eGFR_CRE_. Count and percent or mean and standard deviations are shown. Body mass index was missing for 479 participants (24%), serum albumin was missing for 72 participants (3.6%), and hemoglobin was missing for 46 participants (2.3%). The remaining data was complete. Chronic kidney disease was staged using the eGFR_CRE-CYS_. Abbreviations: eGFR_CRE-CYS_ = estimated Glomerular Filtration Rate using creatinine and cystatin C equation, ACEi/ARB = Angiotensin Converting Enzyme Inhibitor/Angiotensin Receptor Blocker.

|  | Overall | eGFR discrepancy | Reference group |
| --- | --- | --- | --- |
| **Covariates** | N=1988 | N=579 | N=1409 |
| **Age** | 66 (14.1) | 68 (14.3) | 65(14.0) |
| **Female Sex** | 965 (48.5) | 293 (50.6) | 672 (47.7) |
| **Race/Ethnicity** |  |  |  |
| White | 1555 (78.2) | 467 (80.7) | 1088 (77.2) |
| Black | 184 (9.3) | 43 (7.4) | 141 (10.0) |
| Hispanic | 89 (4.5) | 27 (4.7) | 62 (4.4) |
| Asian | 72 (3.6) | 20 (3.5) | 52 (3.7) |
| Other | 88 (4.4) | 22 (3.8) | 66 (4.7) |
| **Body Mass Index (kg/m<sup>2</sup>)** |  |  |  |
| Underweight (<18.5) | 54 (3.6) | 25 (6.1) | 29 (2.6) |
| Normal (18.5 - 24.9) | 442 (29.4) | 126 (30.8) | 316 (28.9) |
| Overweight (25 - 29.9) | 515 (34.2) | 114 (27.9) | 401 (36.6) |
| Obese (≥30) | 493 (32.8) | 144 (35.2) | 349 (31.9) |
| **Comorbidities** |  |  |  |
| Hypertension | 1600 (80.5) | 507 (87.6) | 1093 (77.6) |
| Coronary Artery Disease | 987 (49.6) | 377 (65.1) | 610 (43.3) |
| Diabetes Mellitus | 1008 (50.7) | 393 (67.9) | 615 (43.6) |
| Cirrhosis | 105 (5.3) | 61 (10.5) | 44 (3.1) |
| Human Immunodeficiency Virus | 82 (4.1) | 20 (3.5) | 62 (4.4) |
| Smoking | 806 (40.5) | 266 (45.9) | 540 (38.3) |
| Malnutrition | 275 (13.8) | 97 (16.8) | 178 (12.6) |
| Thyroid disease | 503 (25.3) | 177 (30.6) | 326 (23.1) |
| Chronic kidney disease |  |  |  |
| eGFR 30 - 50 mL/min per 1.72m<sup>2</sup> | 804 (40.4) | 254 (43.9) | 550 (39.0) |
| eGFR < 30 mL/min per 1.73m<sup>2</sup> | 514 (25.8) | 211 (36.5) | 303 (21.5) |
| **Medication Use** |  |  |  |
| ACEi/ARB | 1128 (56.7) | 345 (59.6) | 783 (55.6) |
| Proton Pump Inhibitors | 1291 (64.9) | 441 (76.2) | 850 (60.3) |
| Diuretics | 1282 (64.5) | 474 (81.9) | 808 (57.3) |
| Corticosteroids* | 393 (19.8) | 207 (35.8) | 186 (13.2) |
| **Labs** |  |  |  |
| Serum Creatinine (mg/dL) | 1.62 (1.03) | 1.44 (0.75) | 1.70 (1.12) |
| Serum Cystatin C (mg/L) | 1.82 (0.97) | 2.37 (1.03) | 1.59 (0.84) |
| eGFR<sub>CRE-CYS</sub> (mL/min per 1.73m<sup>2</sup>) | 51 (28) | 41 (22) | 55 (29) |
| Serum Albumin (g/dL) |  |  |  |
| <3.0 | 297 (15.5) | 198 (34.6) | 99 (7.4) |
| 3.0-3.49 | 187 (9.8) | 96 (16.8) | 91 (6.8) |
| 3.5-3.9 | 280 (14.6) | 111 (19.4) | 169 (12.6) |
| ≥4.0 | 1152 (60.1) | 168 (29.3) | 984 (73.3) |
| Blood Urea Nitrogen (mg/dL) |  |  |  |
| ≤17 | 501 (25.2) | 99 (17.1) | 402 (28.6) |
| 17-25 | 526 (26.5) | 107 (18.5) | 419 (29.8) |
| 25-39 | 482 (24.3) | 147 (25.4) | 335 (23.8) |
| >39 | 478 (24.1) | 226 (39.0) | 252 (17.9) |
| Hemoglobin (g/dL) |  |  |  |
| ≤10.0 | 597 (30.0) | 324 (56.0) | 273(19.4) |
| 10 -11.9 | 505 (25.4) | 138 (23.8) | 367 (26.0) |
| ≥12.0 | 886 (44.6) | 117 (20.2) | 769 (54.6) |

Chi-squared or Fisher’s exact tests were used to assess differences in the rate of medication adverse events across groups, as appropriate. As a sensitivity analysis, we performed univariable and multivariable logistic regression to predict the odds of elevated vancomycin level >30 μg/mL; the final multivariable model was adjusted for age, sex, race, baseline eGFR_CRE-CYS_, BMI, diabetes, and corticosteroid use. All comparisons were two-sided, with p<0.05 considered significant. Kaplan-Meier survival curves and multivariable Cox regression models were used to compare 30-day survival across groups. The final multivariable model was adjusted for age, sex, race, baseline eGFR_CRE-CYS_, BMI, coronary artery disease, diabetes, cirrhosis, angiotensin converting enzyme inhibitor or angiotensin receptor blocker use, proton pump inhibitor use, diuretic use, corticosteroid use, serum albumin, and hemoglobin level. All analyses were performed using R 4.1.1 (R Foundation), SAS 9.4 (SAS Institute), and GraphPad PRISM V.9.1.0 (GraphPad Software).

#### Informed Consent

The Massachusetts General Brigham Institutional Review Board approved this retrospective study and waived the need for informed consent.

## Results

### Baseline characteristics

There were 1988 patients with cancer who had a simultaneous creatinine and cystatin C measured between May 4^th^, 2010, and January 26^th^, 2022 (**Figure 1**). Mean age was 66 (SD 14 years), 965 (49%) were female, and 1555 (78%) were non-Hispanic white. Patients with a wide array of cancer types were included (**Supplemental Table 1)**.

**Figure 1.**
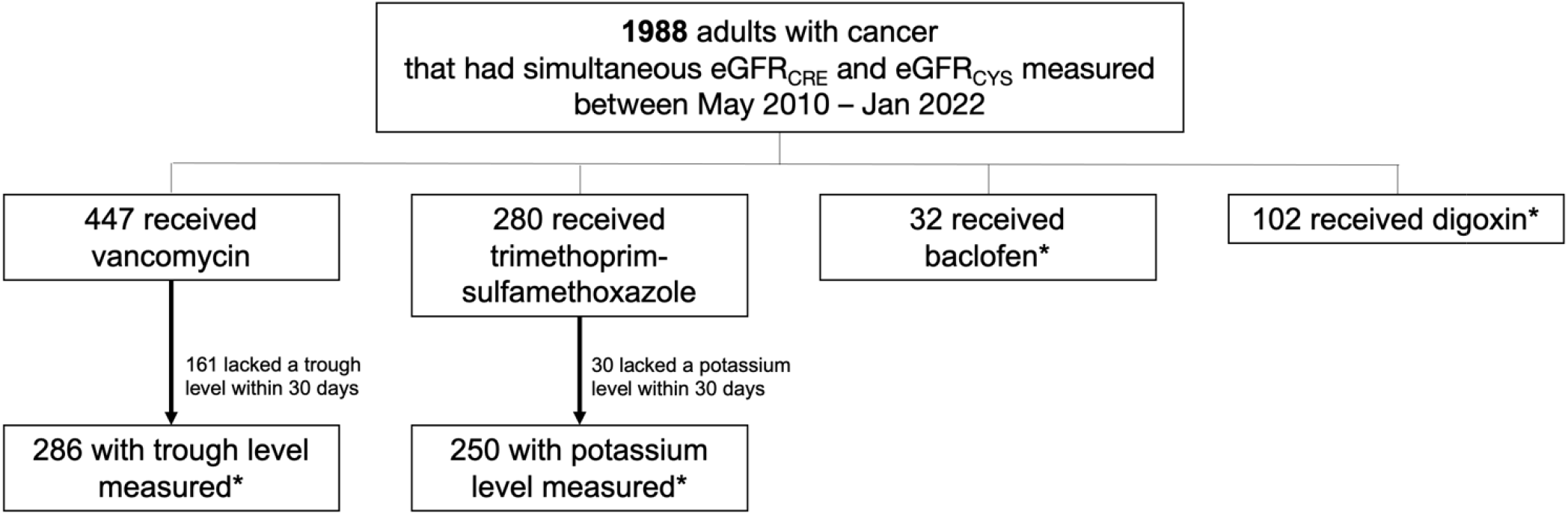
Patient flow. Patient flow. Exposure to each medication was determined by an active prescription within 90 days of the baseline date. *Shows the analyzed sample for each medication. Abbreviations: eGFR_CRE_ = creatinine-based estimated glomerular filtration rate, eGFR_CYS_ = cystatin c-based estimated glomerular filtration rate.

A total of 579 patients (29%) had an eGFR_CYS_ more than 30% lower than eGFR_CRE_. A scatterplot of eGFR_CRE_ vs. eGFR_CYS_ is shown in **Supplemental Figure 1A** and the distribution of the differences between eGFR_CRE_ and eGFR_CYS_ is shown in **Supplemental Figure 1B**. As noted in the methods, the reference group included patients whose eGFR_CYS_ was within 30% of the eGFR_CRE_, as well as the 209 patients (10.5%) whose eGFR_CYS_ was 30% higher than eGFR_CRE_. Predictors of eGFR discrepancy in the multivariable logistic model included white race (adjusted odds ratio [aOR] 1.54, 95% confidence interval [CI] 1.16–2.045, obesity with body-mass-index (BMI) ≥ 30 vs. normal BMI 18.5-24.9 kg/m^2^ (aOR 1.51, 95% CI 1.07–2.13), cirrhosis (aOR 1.82, 95% CI 1.14–2.95), diuretic use (aOR 1.68, 95% CI 1.23–2.30), recent corticosteroid use (aOR 1.70, 95% CI 1.28–2.24), hypoalbuminemia (aOR 5.48, 95% CI 3.84–7.86 for serum albumin < 3.0 vs. ≥4.0 g/dL), and anemia (aOR 2.17, 95% CI 1.55–3.03 for hemoglobin <10.0 vs. ≥ 12.0 g/dL) (**Figure 2, Supplemental Table 2**).

**Figure 2.**
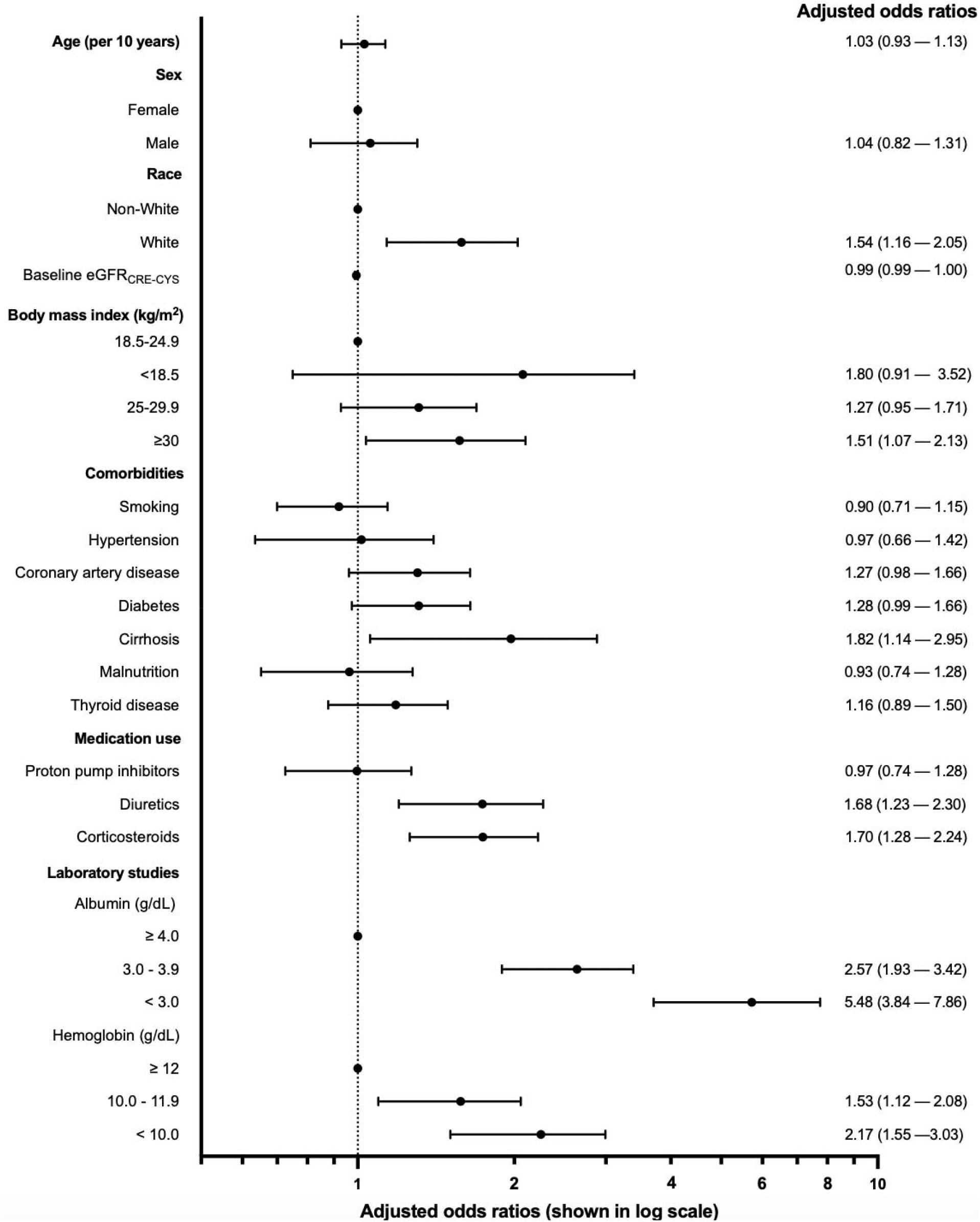
Predictors of eGFR discrepancy. **Forrest plot showing adjusted odds ratios.** Logistic regression models were used to estimate the association between baseline characteristics and the eGFR discrepancy (eGFR_CYS_ > 30% lower than eGFR_CRE_) in patients with cancer. The unadjusted and adjusted models are also shown in **Supplemental Table 2**. Abbreviations: eGFR_CRE-CYS_ = estimated Glomerular Filtration Rate calculated using the 2021 race-free combined serum creatinine and cystatin C equation

Hypoalbuminemia and anemia were the baseline factors most strongly associated with having an eGFR discrepancy; there was a stepwise increase in the likelihood of eGFR discrepancy as albumin and hemoglobin levels decreased (**Figure 3, Supplemental Figure 2**). Among patients with albumin ≥ 4.0 g/dL only 174/1224 (14%) had an eGFR discrepancy, compared to 198/297 (67%) in patients with albumin < 3.0 g/dL. Among patients with hemoglobin ≥ 12 g/dL only 117/886 (13%) had an eGFR discrepancy, compared to 130/196 (66%) in patients with hemoglobin < 8.0 g/dL.

**Figure 3.**
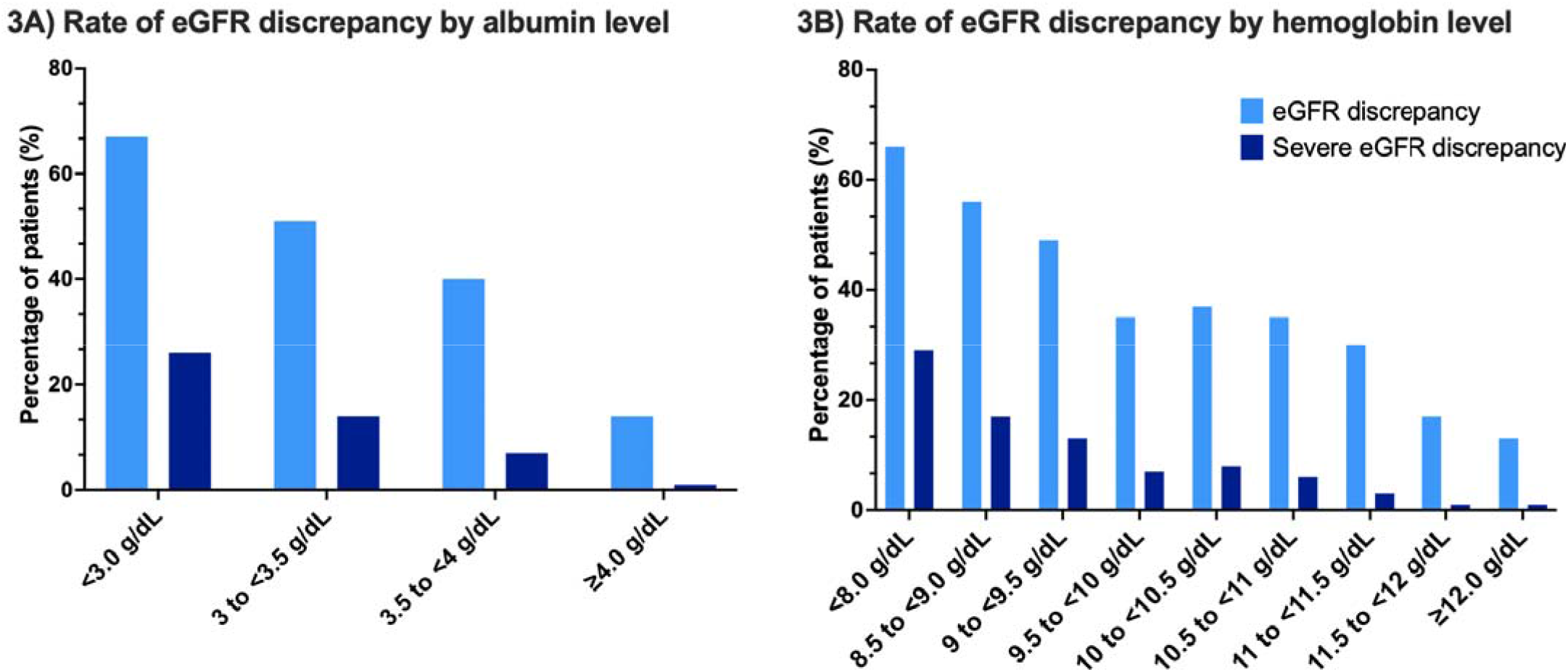
Rate of eGFR discrepancy by albumin and hemoglobin levels. eGFR discrepancy defined by eGFR_CYS_ > 30% lower than eGFR_CRE_ shown with light blue bars, and severe eGFR discrepancy (eGFR_CYS_ > 50% lower than eGFR_CRE_ and eGFR_CYS_ < 30mL/min/1.73m^2^) shown with dark blue bars become more common in patients with worsening hypoalbuminemia (**3A**) and anemia (**3B**).

There were 139 patients (7.0% of the overall cohort) who had severe eGFR discrepancy (eGFR_CYS_ > 50% lower than eGFR_CRE_ and eGFR_CYS_ < 30mL/min/1.73m^2^) (**Supplemental Table 3**). Predictors of severe eGFR discrepancy were similar to eGFR discrepancy and are shown in **Supplemental Table 4**. The rate of eGFR discrepancy and severe eGFR discrepancy varied by cancer type (**Supplemental Figure 3**).

### Medication-related Adverse Events

#### Vancomycin

There were 447 patients who received vancomycin within 90 days of the baseline date, of whom 286 (64%) had a vancomycin trough measured (**Figure 1**). Patients with eGFR discrepancy were more likely to have significantly elevated vancomycin trough levels than the reference group: 129 of 193 (67%) vs. 37 of 93 (40%) of the reference group had a vancomycin level above the therapeutic range (*P* < 0.001); 46 of 193 (24%) vs. 9 of 93 (10%) had trough level >30 μcg/mL (*P* = 0.004); 15 of 193 (8%) vs. 0 of 93 (0%) had a trough level >40 μcg/mL (*P* = 0.003) (**Figure 4A**). The rate of elevated vancomycin trough levels was even higher in patients with severe eGFR discrepancy (**Figure 4A)**. After adjustment for baseline demographics, comorbidities, and baseline laboratory studies, patients with eGFR discrepancy had a 2.30-fold adjusted OR (95% CI 1.05 – 5.51) of having a significantly elevated vancomycin trough level >30 μg/mL (**Supplemental Table 5**).

**Figure 4.**
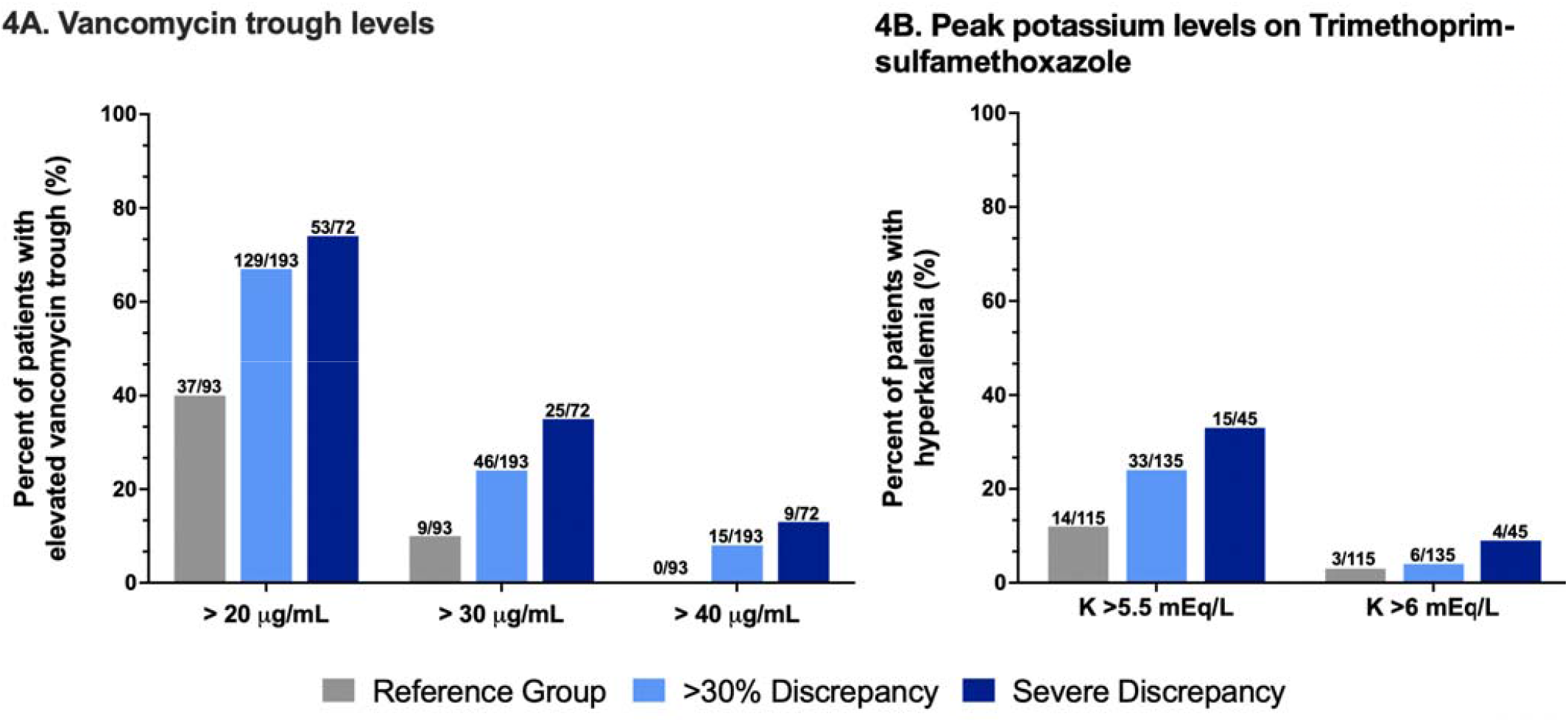
Rate of supratherapeutic vancomycin levels and trimethoprim-sulfamethoxazole-related hyperkalemia in patients with eGFR discrepancy. **Figure 4A**. Highest trough vancomycin levels obtained within 30 days of starting vancomycin in patients with eGFR discrepancy (eGFR_CYS_ more than 30% lower than eGFR_CRE_), severe eGFR discrepancy (eGFR_CYS_ more than 50% lower than eGFR_CRE_ and eGFR_CYS_ < 30mL/min/1.73m^2^), and the reference group. We excluded any vancomycin level obtained less than 6 hours after the last administered vancomycin dose. **Figure 4B**. Rate of Grade 2 or 3 hyperkalemia in patients receiving trimethoprim-sulfamethoxazole.

### Trimethoprim-sulfamethoxazole

There were 280 patients who received trimethoprim-sulfamethoxazole within 90 days of the baseline date. We excluded 30 (11%) who did not have a serum potassium level checked within 30 days of starting trimethoprim-sulfamethoxazole (**Figure 1**). Patients with eGFR discrepancy were more likely to experience hyperkalemia (potassium >5.5mEq/L) after starting trimethoprim-sulfamethoxazole compared to the reference group 33 of 135 (24%) vs. 14 of 115 (12%), *P* = 0.013 (**Figure 4B**). The rate of trimethoprim-sulfamethoxazole-related hyperkalemia was even greater in patients with severe eGFR discrepancy, affecting 15 of 45 (33%) of patients (*P* = 0.0018) (**Figure 4B**). A similar trend was found when evaluating the rate of grade 3 hyperkalemia (defined by a potassium level > 6.0mEq) (**Figure 4B**).

### Baclofen

There were 32 patients newly prescribed baclofen within 90 days of baseline (**Figure 1**). Five of the 20 patients (25%) with eGFR discrepancy developed clinical evidence of baclofen toxicity which prompted discontinuation of the medication compared to none of the 12 patients in the reference group (*P* = 0.13) (**Figure 5A**). Among those with severe eGFR discrepancy, 3 out of 8 (37.5%) developed baclofen toxicity. The most common symptom of baclofen toxicity was somnolence/depressed level of consciousness (3 cases). Two additional patients developed severe orthostatic hypotension.

**Figure 5.**
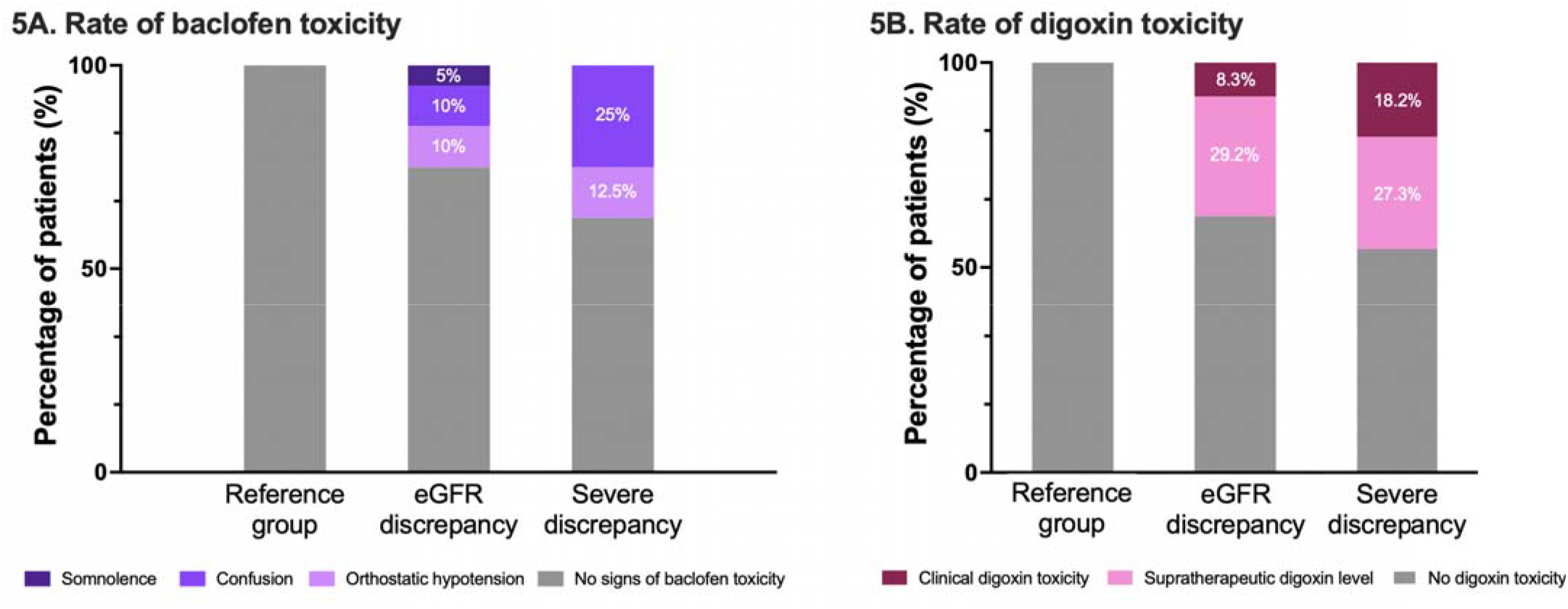
Rate of baclofen toxicity and digoxin toxicity in patients with eGFR discrepancy. **Figure 5A**. Rate of baclofen toxicity in patients with eGFR discrepancy, severe eGFR, discrepancy and the reference group. **Figure 5B**. Rate of supratherapeutic digoxin level defined by >2.0μg/dL. Both cases of clinical digoxin toxicity occurred in patients with severe eGFR discrepancy (eGFR_CYS_ more than 50% lower than eGFR_CRE_ and eGFR_CYS_ < 30mL/min/1.73m^2^). There were no cases of supratherapeutic digoxin levels or digoxin toxicity in the reference group.

### Digoxin

There were 102 patients who were prescribed digoxin (**Figure 1**), of whom 34 (33%) had at least one digoxin level measured. Out of the 24 patients with eGFR discrepancy, 9 patients (38%) had a digoxin trough level above the therapeutic range (>2.0 ng/mL) compared to none of the 10 patients in the reference group (*P* = 0.034). Two patients (8.3%) were diagnosed with clinical digoxin toxicity, including one who required digoxin immune fab (**Supplemental Table 6**); both patients diagnosed with clinical digoxin toxicity met criteria for severe eGFR discrepancy (**Figure 5B**).

#### Thirty-day survival

132 patients (7%) died within 30 days and 173 (9%) were lost to follow-up prior to 30 days. There was significantly higher 30-day mortality in patients with eGFR discrepancy compared to the reference group (**Figure 6)**. Even after adjustment for age, sex, race, baseline comorbidities, laboratory tests, and medication use, patients with eGFR discrepancy had a 1.97-fold increased hazard of death (95% CI 1.29–3.01), compared to the reference group (**Figure 6, Supplemental Table 7**).

**Figure 6.**
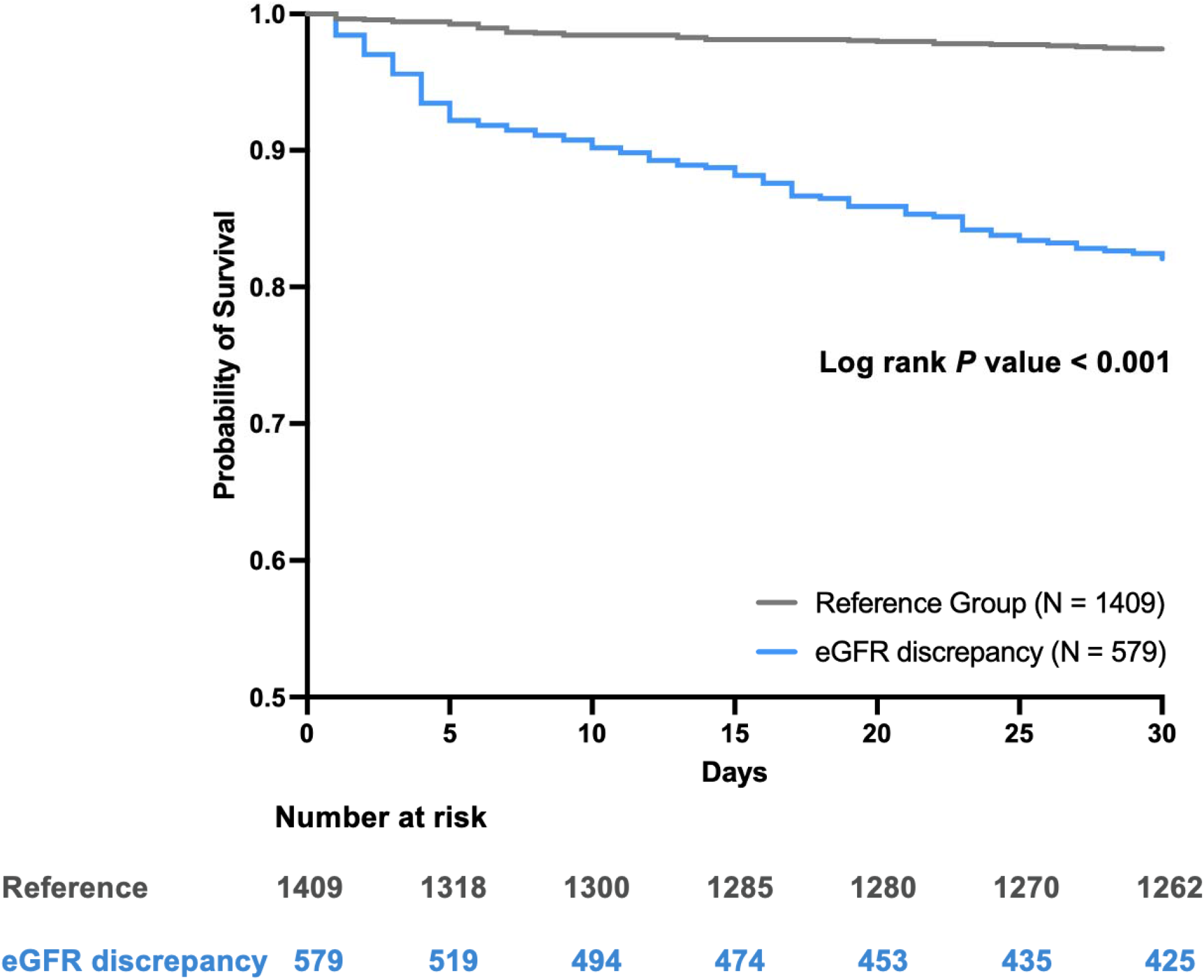
Kaplan-Meier curve of 30-day survival. Survival analysis using Kaplan Meier method comparing 30-day survival between reference group and those who had eGFR discrepancy. Unadjusted and adjusted model is shown in Supplemental Table 7.

## Discussion

Our study showed that in a cohort of patients with a history of cancer who have concurrent creatinine and cystatin C measurement, almost 1 out of 3 had an eGFR_CYS_ more than 30% lower than eGFR_CRE_. The high rate of eGFR discrepancy in patients with cancer poses a challenge for clinical decision making and signifies an important knowledge gap in appropriate dose adjustment of medications that primarily undergo renal clearance. We found a considerably higher rate of adverse events associated with select renally-cleared medications in patients with eGFR discrepancy compared to our reference group.

Accurate dosing of renally-cleared medications is a challenge in patients with cancer, among whom sarcopenia is common and overestimation of GFR by creatinine-based equations has been a major concern in clinical practice.^10, 26-28^ Cystatin C, which is produced by all nucleated cells and is not dependent on diet or muscle mass, has been validated as an alternative marker to estimate kidney function. However, Cystatin C levels may be falsely increased in patients with obesity, inflammation, current smoking, and corticosteroid therapy.^29-32^ In 2021, the National Kidney Foundation and the American Society of Nephrology Task Force recommended that clinicians estimate GFR using a combined equation incorporating both cystatin C and serum creatinine.^33^ A recent study of 1200 patients with solid tumors who underwent measured GFR found that eGFR_CRE_ overestimated measured GFR, eGFR_CYS_ underestimated measured GFR, and that the most accurate and precise eGFR was obtained using the combined equation incorporating both cystatin C and serum creatinine.^9^ We note that risk factors for eGFR underestimation with cystatin C is greater in patients with higher BMI, current and former smokers, low albumin, higher C-reactive protein, and metastatic disease. It is important to note the overlap with the predictors of eGFR discrepancy we identified in this study. Because we lacked measured GFR, we are unable to determine the accuracy of eGFR_CRE_ and eGFR_CYS_, however, we found that when a large eGFR discrepancy exists, patients with cancer are at higher risk of adverse events from renally-cleared medications.

Our study demonstrated a higher rate of supratherapeutic vancomycin levels in patients with eGFR discrepancy. Vancomycin is a very commonly used intravenous antibiotic in hospitalized patients that is predominantly eliminated by the kidney (>90%). Several studies have demonstrated that vancomycin clearance and target trough achievement may be more accurately predicted by eGFR_CYS_ than eGFR_CRE_.^34-38^ A previously published quality improvement initiative that included 399 patients found that a vancomycin dosing algorithm using eGFR estimated by both creatinine and cystatin C (N = 135) was more likely to achieve therapeutic vancomycin trough levels (50% vs. 28%, p< 0.001) compared to an algorithm using eGFR_CRE_ alone (N = 264).^36^ Trimethoprim-sulfamethoxazole is another commonly used antibiotic in the inpatient and outpatient setting. Trimethoprim can have an “amiloride-like” effect by inhibiting potassium secretion in the distal convoluted tubule; patients with impaired kidney function are much more likely to develop clinically significant hyperkalemia when treated with trimethoprim-sulfamethoxazole.^39^ Hyperkalemia occurred more commonly in patients with eGFR discrepancy compared to the reference group. Digoxin is a cardiac glycoside medication approved to treat atrial fibrillation and congestive heart failure that has a narrow therapeutic index. Digoxin is cleared by the kidneys and its toxicity is dose dependent. There have been conflicting reports regarding use of creatinine versus cystatin C to predict digoxin clearance.^40-44^ Here, we found that patients with eGFR discrepancy were significantly more likely to have supratherapeutic digoxin trough levels compared to the reference group, and both cases of symptomatic digoxin toxicity occurred in patients with eGFR discrepancy. Baclofen is a muscle relaxant that is commonly prescribed in patients with cancer to inhibit the hiccup reflex. Baclofen is primarily eliminated by the kidney. In patients with impaired kidney function, baclofen accumulation can occur after just a single dose, and can lead to profound central nervous system suppression, ranging from encephalopathy, coma, areflexia, hypotension, and cardiac arrest.^45, 46^ Our study showed that symptomatic baclofen toxicity is common in patients with eGFR discrepancy (affecting 25% of patients), whereas no events occurred in the reference group, suggesting that the eGFR_CYS_ should be considered when prescribing baclofen to patients with cancer. To the best of our knowledge, this is the first study to evaluate eGFR_CYS_ and eGFR_CRE_ in patients receiving baclofen. Taken together, our findings suggest that relying on creatinine-based eGFR alone for medication dosing may be inadequate in patients with cancer and highlights the need to consider eGFR_CYS_ as well.

Finally, consistent with prior knowledge that incorporation of cystatin C adds precision to the eGFR equation in patients with malnutrition, sarcopenia, and cirrhosis,^9, 47, 48^ we note that hypoalbuminemia and anemia are important predictors of eGFR discrepancy in patients with cancer. It is likely that these laboratory abnormalities that signify chronic illness are associated with cancer-related sarcopenia. Future prospective studies that include measured GFR are needed to validate this finding. In addition, we found that eGFR discrepancy is associated with significantly higher 30-day mortality even after adjustment for demographics, comorbidities, baseline laboratory studies, and medication use. Although higher serum creatinine and cystatin C have each been shown to be associated with increased mortality in multiple clinical settings,^49-54^ our finding suggests that discrepancy between creatinine and cystatin C, in addition to the absolute values of either marker, adds further information and potentially serves as an independent predictor of death.

Our study has several limitations. First, cystatin C was only available on select patients, where it has been ordered as a part of routine care. Since cystatin C is not routinely used in clinical practice, our population was likely enriched for patients in whom clinicians suspected an eGFR discrepancy or kidney injury might exist, this likely an overestimate of the rate of eGFR discrepancy in the oncology population in general. However, the selection bias should be balanced between the eGFR discrepancy group and the reference group, which preserves the validity of comparison of medication-related adverse events between the two groups. Second, we only used a one-time assessment of creatinine and cystatin C, which may not reflect a steady state at the time of measurement. Third, we were not able to determine cancer stage from our dataset, which may be an important non-GFR determinant of cystatin C and creatinine.^55, 56^ Fourth, it is possible that clinician knowledge of the eGFR_CYS_ could have influenced medication dosing; however, such practice would have biased our results toward the null. Accordingly, the magnitude of association between eGFR discrepancy and medication-related adverse events that we report here is likely an underestimate. Finally, our study lacks gold standard GFR measurement given the retrospective design; however, comparing adverse outcomes of renally-dosed medications serves as surrogate marker for accuracy of eGFR.

In conclusion, we found that a >30% eGFR discrepancy is common in patients with cancer and is associated with an increase in adverse events related to commonly used, renally-cleared medications. Future prospective studies are needed to improve and personalize the approach to GFR estimation and medication dosing in patients with cancer.

## Data Availability

All data produced in the present study are available upon reasonable request to the authors

## Disclaimers

The results presented in this report have been presented at the American Society of Onconephrology Symposium in a poster format, however they have not been published previously in whole or part.

---

**Supplemental Figure 1.**
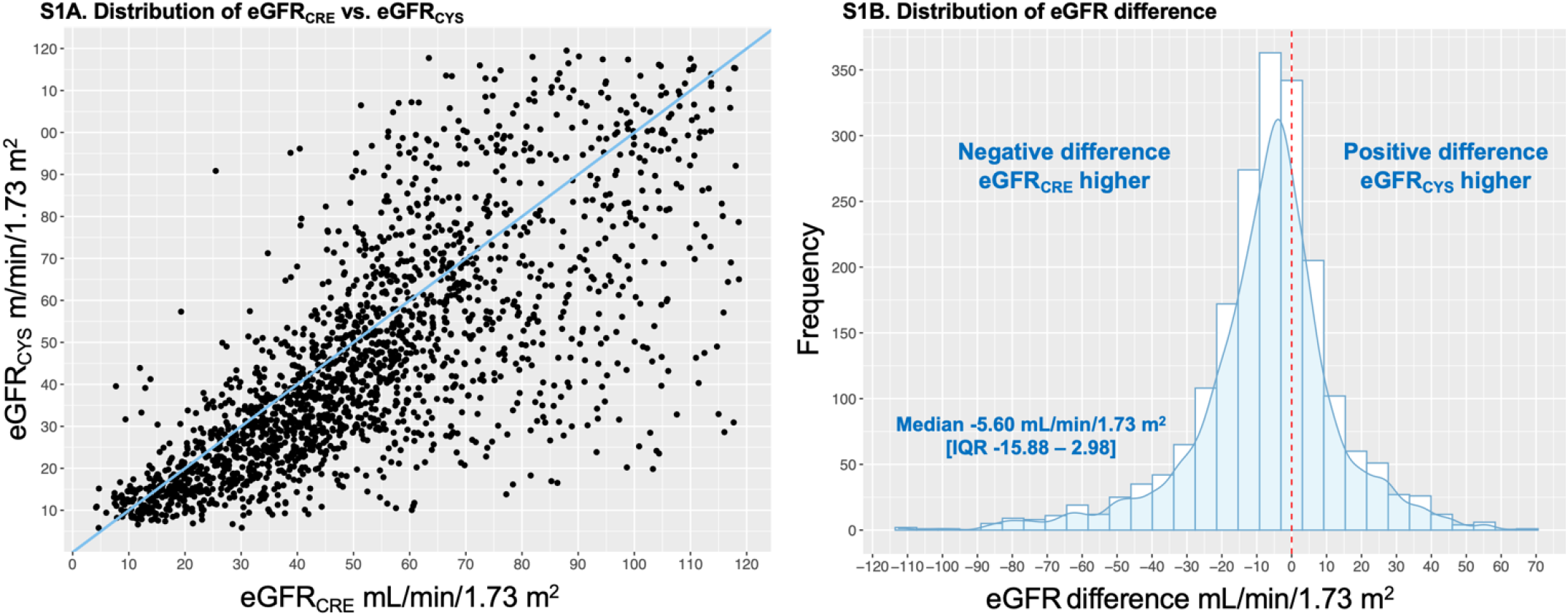
Scatter plot of creatinine-based and cystatin C-based eGFR, and distribution of eGFR difference. Scatterplot showing distribution of eGFR_CRE_ and eGFR_CYS_ among patients with cancer (S1A); the blue line is the line of equality. Figure S1B. A histogram and superimposed density curve showing the distribution of eGFR difference defined by eGFR_CYS_ minus eGFR_CRE_. The red dotted line signifies equivalence between eGFR_CRE_ and eGFR_CYS_. <u>Abbreviations</u>: IQR, interquartile range

**Supplemental Figure 2.**
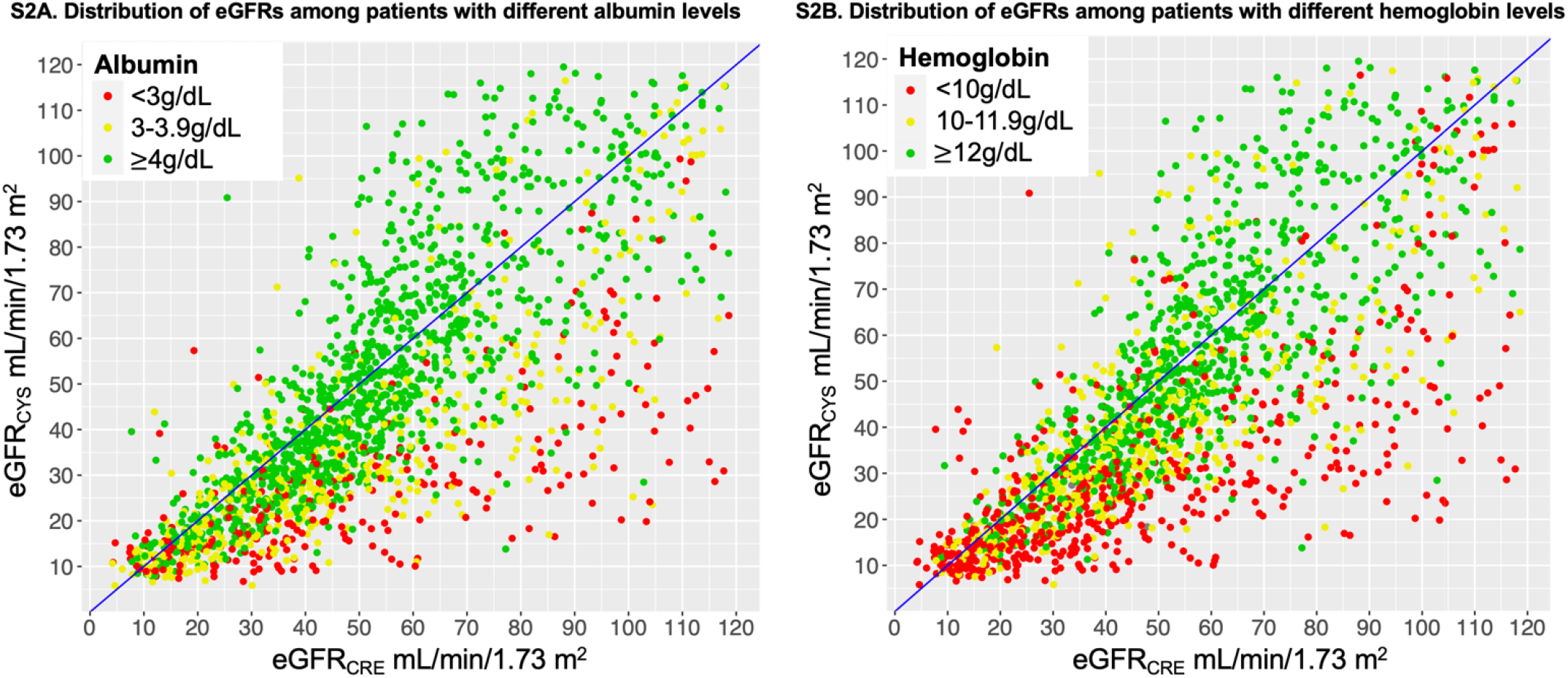
Scatter plot of creatinine-based and cystatin C-based eGFR by albumin and hemoglobin levels. Scatterplot showing distribution of eGFRs among patients with cancer stratified by different albumin levels (S2A) and hemoglobin levels (S2B) as shown. The identity line (line of equality) is shown in blue.

**Supplemental Figure 3.**
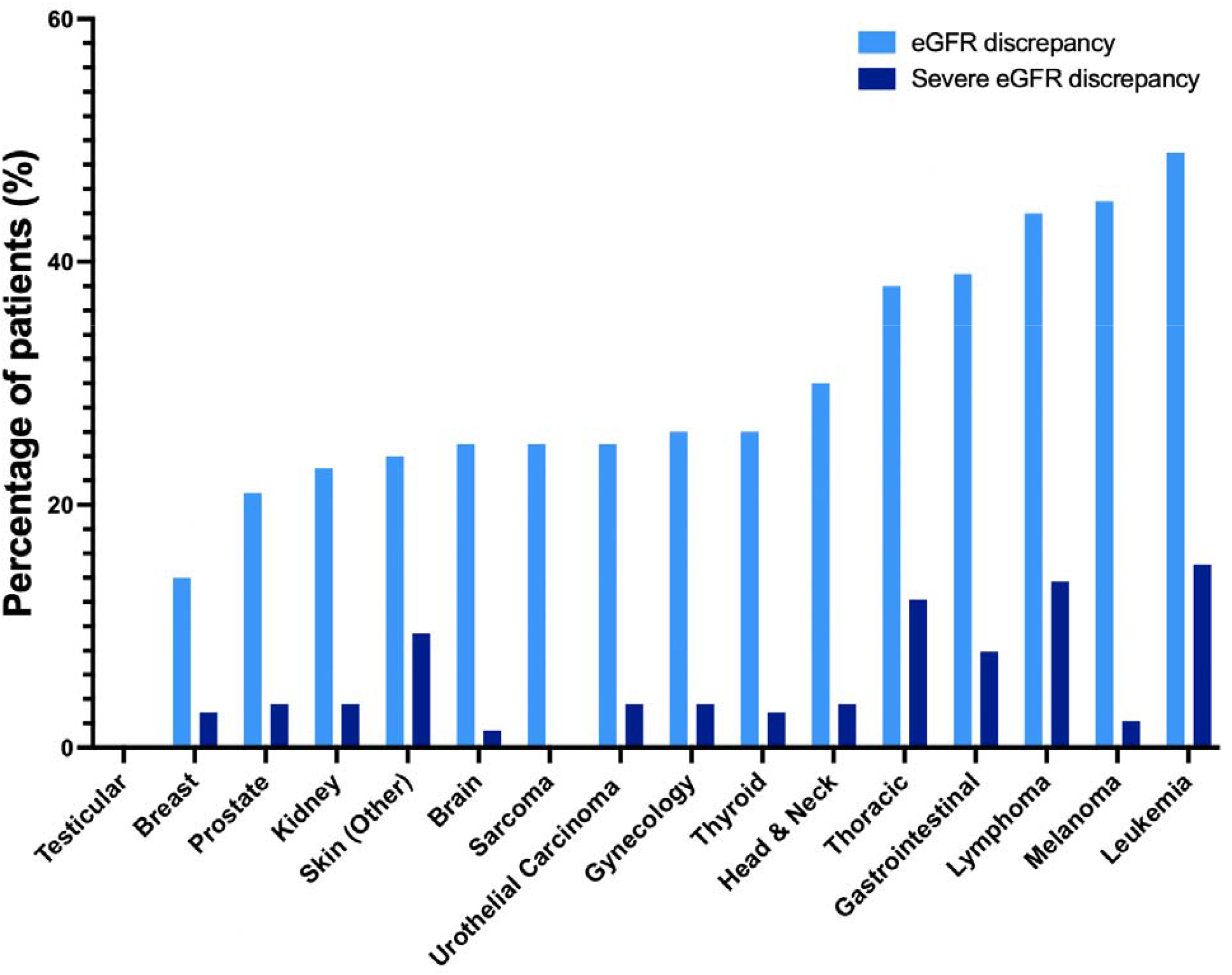
Rate of eGFR discrepancies by cancer type Rate of eGFR discrepancy and severe eGFR discrepancy by cancer type. eGFR discrepancy is defined by an eGFR_CYS_ more than 30% lower than eGFR_CRE_. Severe eGFR discrepancy was defined as an eGFR_CYS_ more than 50% lower than eGFR_CRE_ and eGFR_CYS_ < 30mL/min/1.73m^2^.

**Supplemental Figure 4.**
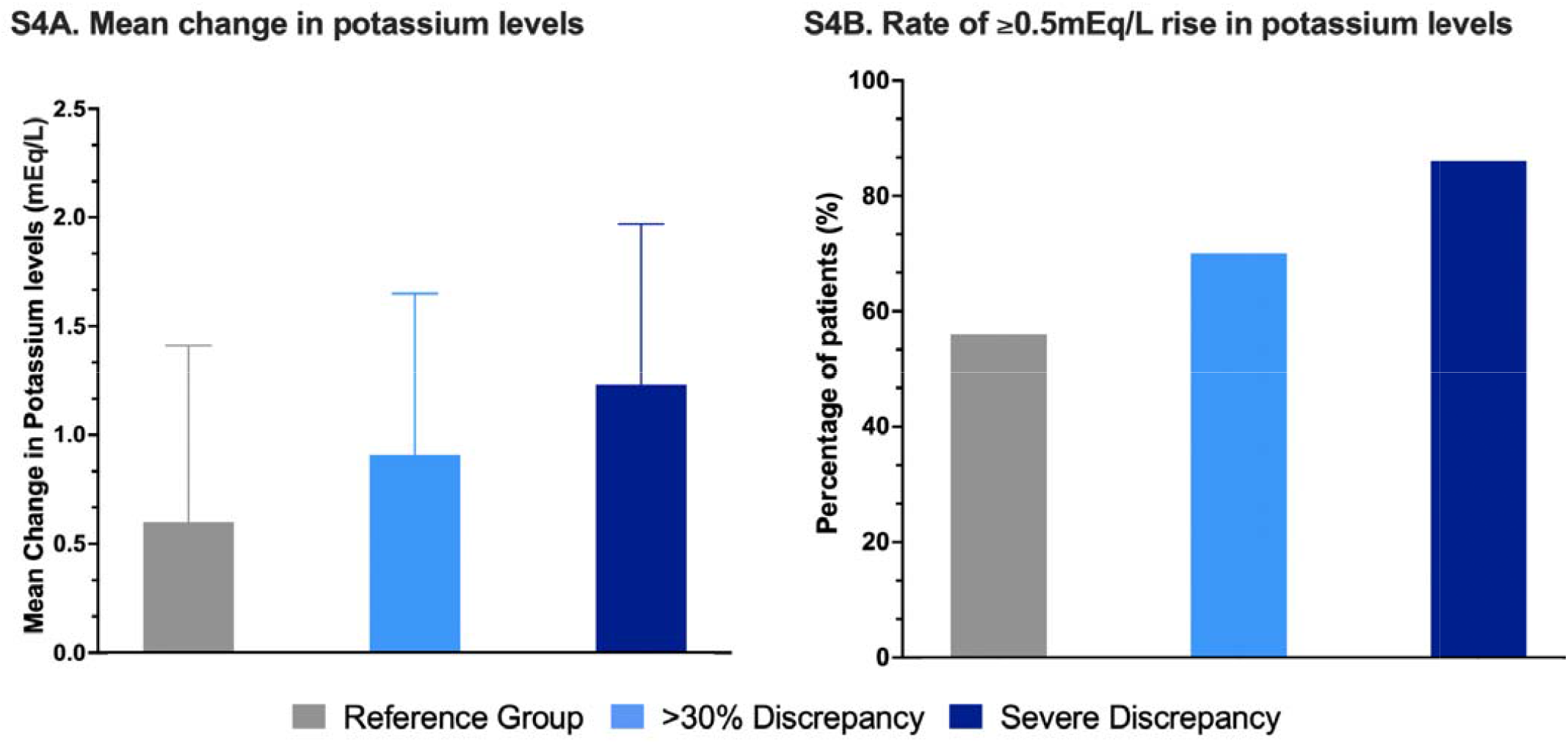
Changes in potassium levels among patients who received trimethoprim-sulfamethoxazole. **Supplemental Figure S4A**. Mean change in potassium levels with standard deviations among trimethoprim-sulfamethoxazole recipients; error bars show standard deviation. **S4B**. Rate of ≥0.5mEq/L rise in potassium levels among trimethoprim-sulfamethoxazole recipients. eGFR discrepancy is defined by an eGFR_CYS_ more than 30% lower than eGFR_CRE_. Severe eGFR discrepancy was defined as an eGFR_CYS_ more than 50% lower than eGFR_CRE_ and eGFR_CYS_ < 30mL/min/1.73m^2^.

**Supplemental Table 1.** Cancer type. Cancer type was determined by the most commonly appearing cancer-related diagnosis code prior to the baseline date. eGFR discrepancy is defined by an eGFR_CYS_ more than 30% lower than eGFR_CRE_.

|  | Overall | eGFR discrepancy | Reference group |
| --- | --- | --- | --- |
|  | N=1988 | N=579 | N=1409 |
| **Cancer types\*** |  |  |  |
| Urothelial | 92 (4.6) | 23 (4.0) | 69 (4.9) |
| Brain | 40 (2.0) | 10 (1.7) | 30 (2.1) |
| Breast | 228 (11.5) | 32 (5.5) | 196 (13.9) |
| Gastrointestinal | 177 (8.9) | 69 (11.9) | 108 (7.7) |
| Gynecologic | 141 (7.1) | 36 (6.2) | 105 (7.5) |
| Head & Neck | 50 (2.5) | 15 (2.6) | 35 (2.5) |
| Leukemia | 129 (6.5) | 63 (10.9) | 66 (4.7) |
| Lymphoma | 123 (6.2) | 54 (9.3) | 69 (4.9) |
| Melanoma | 29 (1.5) | 13 (2.2) | 16 (1.1) |
| Prostate | 147 (7.4) | 31 (5.4) | 116 (8.2) |
| Kidney | 162 (8.1) | 37 (6.4) | 125 (8.9) |
| Sarcoma | 4 (0.2) | 1 (0.2) | 3 (0.2) |
| Testicular | 1 (0.1) | 0 (0.0) | 1 (0.1) |
| Thoracic | 114 (5.7) | 43 (7.4) | 71 (5.0) |
| Thyroid | 43 (2.2) | 11 (1.9) | 32 (2.3) |
| Skin (Other) | 298 (15.0) | 71 (12.3) | 227 (16.1) |

**Supplemental Table 2.**
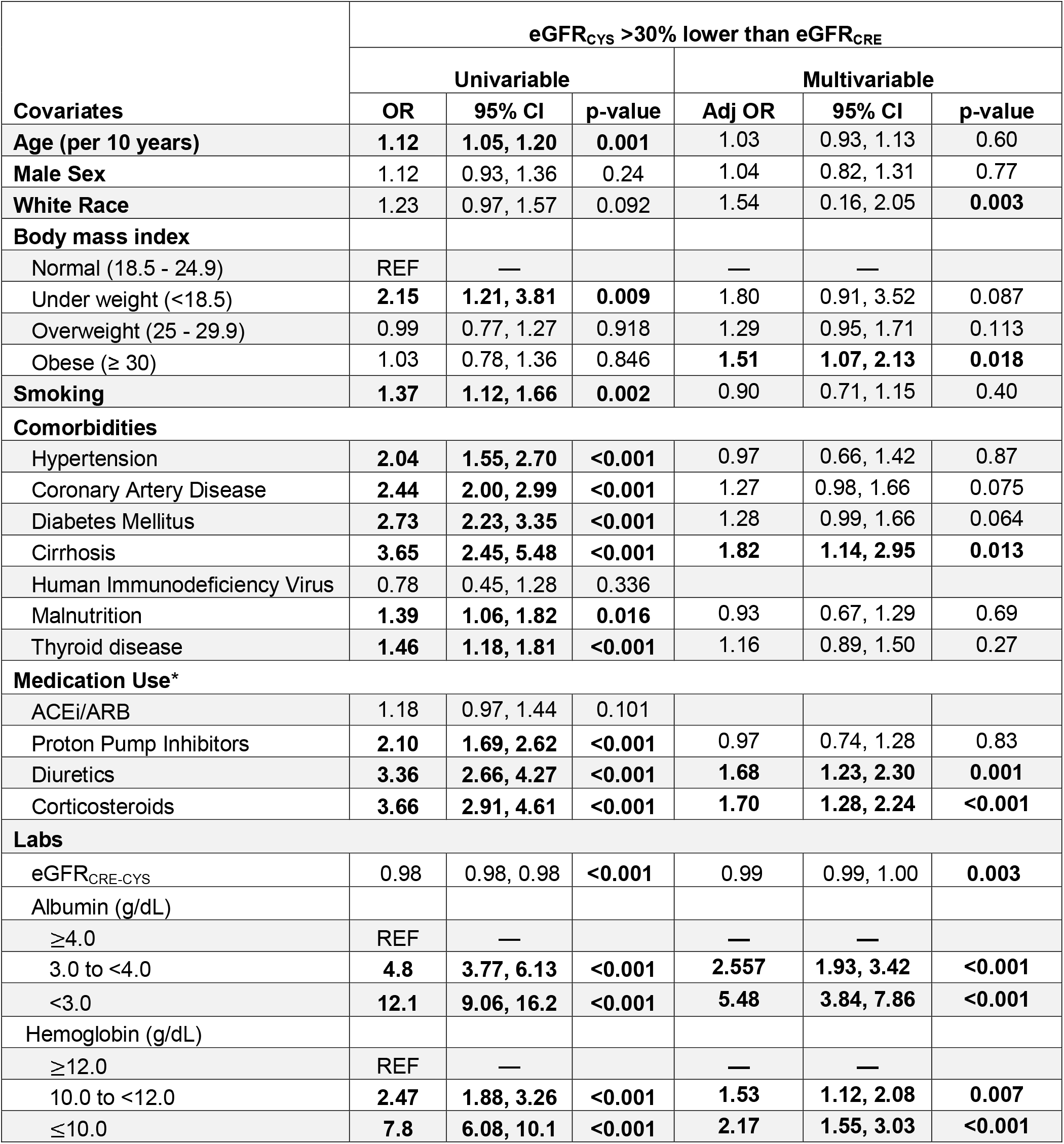
Predictors of eGFR_CYS_ more than 30% lower than eGFR_CRE_. Univariable and multivariable logistic regression model. *Chronic medication use was defined within 1 year prior to baseline; corticosteroid use was defined within 30 days of baseline. In all cases the population median was imputed for missing variables (Body mass index was missing for 479 participants, serum albumin was missing for 72 participants, hemoglobin was missing for 46 participants). Abbreviations eGFR_CRE-CYS_ = estimated Glomerular Filtration Rate calculated using the 2021 race-free combined serum creatinine and cystatin C equation, ACEi/ARB = Angiotensin Converting Enzyme Inhibitor/Angiotensin Receptor Blocker.

| Covariates | eGFR<sub>CYS</sub> >30% lower than eGFR<sub>CRE</sub> |  |  |  |  |  |
| --- | --- | --- | --- | --- | --- | --- |
|  | Univariable |  |  | Multivariable |  |  |
|  | OR | 95% CI | p-value | Adj OR | 95% CI | p-value |
| **Age (per 10 years)** | **1.12** | **1.05, 1.20** | **0.001** | 1.03 | 0.93, 1.13 | 0.60 |
| **Male Sex** | 1.12 | 0.93, 1.36 | 0.24 | 1.04 | 0.82, 1.31 | 0.77 |
| **White Race** | 1.23 | 0.97, 1.57 | 0.092 | 1.54 | 0.16, 2.05 | **0.003** |
| **Body mass index** |  |  |  |  |  |  |
| Normal (18.5 - 24.9) | REF | — |  | — | — |  |
| Under weight (<18.5) | **2.15** | **1.21, 3.81** | **0.009** | 1.80 | 0.91, 3.52 | 0.087 |
| Overweight (25 - 29.9) | 0.99 | 0.77, 1.27 | 0.918 | 1.29 | 0.95, 1.71 | 0.113 |
| Obese (≥ 30) | 1.03 | 0.78, 1.36 | 0.846 | **1.51** | **1.07, 2.13** | **0.018** |
| **Smoking** | **1.37** | **1.12, 1.66** | **0.002** | 0.90 | 0.71, 1.15 | 0.40 |
| **Comorbidities** |  |  |  |  |  |  |
| Hypertension | **2.04** | **1.55, 2.70** | **<0.001** | 0.97 | 0.66, 1.42 | 0.87 |
| Coronary Artery Disease | **2.44** | **2.00, 2.99** | **<0.001** | 1.27 | 0.98, 1.66 | 0.075 |
| Diabetes Mellitus | **2.73** | **2.23, 3.35** | **<0.001** | 1.28 | 0.99, 1.66 | 0.064 |
| Cirrhosis | **3.65** | **2.45, 5.48** | **<0.001** | **1.82** | **1.14, 2.95** | **0.013** |
| Human Immunodeficiency Virus | 0.78 | 0.45, 1.28 | 0.336 |  |  |  |
| Malnutrition | **1.39** | **1.06, 1.82** | **0.016** | 0.93 | 0.67, 1.29 | 0.69 |
| Thyroid disease | **1.46** | **1.18, 1.81** | **<0.001** | 1.16 | 0.89, 1.50 | 0.27 |
| **Medication Use\*** |  |  |  |  |  |  |
| ACEi/ARB | 1.18 | 0.97, 1.44 | 0.101 |  |  |  |
| Proton Pump Inhibitors | **2.10** | **1.69, 2.62** | **<0.001** | 0.97 | 0.74, 1.28 | 0.83 |
| Diuretics | **3.36** | **2.66, 4.27** | **<0.001** | **1.68** | **1.23, 2.30** | **0.001** |
| Corticosteroids | **3.66** | **2.91, 4.61** | **<0.001** | **1.70** | **1.28, 2.24** | **<0.001** |
| **Labs** |  |  |  |  |  |  |
| eGFR<sub>CRE-CYS</sub> | 0.98 | 0.98, 0.98 | **<0.001** | 0.99 | 0.99, 1.00 | **0.003** |
| Albumin (g/dL) |  |  |  |  |  |  |
| ≥4.0 | REF | — |  | — | — |  |
| 3.0 to <4.0 | **4.8** | **3.77, 6.13** | **<0.001** | **2.557** | **1.93, 3.42** | **<0.001** |
| <3.0 | **12.1** | **9.06, 16.2** | **<0.001** | **5.48** | **3.84, 7.86** | **<0.001** |
| Hemoglobin (g/dL) |  |  |  |  |  |  |
| ≥12.0 | REF | — |  | — | — |  |
| 10.0 to <12.0 | **2.47** | **1.88, 3.26** | **<0.001** | **1.53** | **1.12, 2.08** | **0.007** |
| ≤10.0 | **7.8** | **6.08, 10.1** | **<0.001** | **2.17** | **1.55, 3.03** | **<0.001** |

**Supplemental Table 3.** Characteristics of patients with severe eGFR discrepancy. Severe eGFR discrepancy was defined as eGFR_CYS_ more than 50% lower than eGFR_CRE_ and eGFR_CYS_ < 30mL/min/1.73m^2^. Count and percent or mean and standard deviations are shown. Body mass index was missing for 479 participants, serum albumin was missing for 72 participants, hemoglobin was missing for 46 participants. The remaining data was complete. The cohort median was imputed for missing data. Cancer type was determined by the most common cancer-related diagnosis code appearing prior to the baseline date. Abbreviations: eGFR = estimated glomerular filtration rate, ACEi/ARB = Angiotensin Converting Enzyme Inhibitor/Angiotensin Receptor Blocker.

|  | Overall | Severe eGFR discrepancy |
| --- | --- | --- |
| **Covariates** | N=1988 | N=139 |
| **Age** | 66 (14) | 68 (13) |
| **Female Sex** | 965 (48.5) | 60 (43.2) |
| **Race/Ethnicity** |  |  |
| White | 1555 (78.2) | 114 (82.0) |
| Black | 184 (9.3) | 10 (7.2) |
| Hispanic | 89 (4.5) | 6 (4.3) |
| Asian | 72 (3.6) | 4 (2.9) |
| Other | 88 (4.4) | 5 (3.6) |
| **Body Mass Index (kg/m<sup>2</sup>)** |  |  |
| Underweight (<18.5) | 54 (3.6) | 4 (4.6) |
| Normal (18.5 - 24.9) | 442 (29.4) | 28 (32.2) |
| Overweight (25 - 29.9) | 515 (34.2) | 26 (29.9) |
| Obese (≥30) | 493 (32.8) | 29 (33.3) |
| **Comorbidities** |  |  |
| Hypertension | 1600 (80.5) | 124 (89.2) |
| Coronary Artery Disease | 987 (49.6) | 99 (71.2) |
| Diabetes Mellitus | 1008 (50.7) | 112 (80.6) |
| Cirrhosis | 105 (5.3) | 20 (14.4) |
| Human Immunodeficiency Virus | 82 (4.1) | 6 (4.3) |
| Smoking | 806 (40.5) | 67 (48.2) |
| Malnutrition | 275 (13.8) | 21 (15.1) |
| **Medication Use** |  |  |
| ACEi/ARB | 1128 (56.7) | 77 (55.4) |
| Proton Pump Inhibitors | 1291 (64.9) | 115 (82.7) |
| Diuretics | 1282 (64.5) | 122 (87.8) |
| **Labs** |  |  |
| Serum Creatinine (mg/dL) | 1.62 (1.03) | 1.56 (0.65) |
| Serum Albumin (g/dL) |  |  |
| <3.0 | 297 (15.5) | 77 (55.4) |
| 3.0-3.49 | 187 (9.8) | 27 (19.4) |
| 3.5-3.9 | 280 (14.6) | 20 (14.4) |
| ≥4.0 | 1152 (60.1) | 15 (10.8) |
| Blood Urea Nitrogen (mg/dL) |  |  |
| ≤17 | 501 (25.2) | 4 (2.9) |
| 17-25 | 526 (26.5) | 6 (4.3) |
| 25-39 | 482 (24.3) | 30 (21.6) |
| >39 | 478 (24.1) | 99 (71.2) |
| Hemoglobin (g/dL) |  |  |
| ≤10.0 | 597 (30.0) | 111 (79.9) |
| 10.0-11.9 | 505 (25.4) | 18 (12.9) |
| ≥12.0 | 886 (44.6) | 10 (7.2) |
| **Cancer types** |  |  |
| Urothelial | 92 (4.6) | 5 (3.6) |
| Brain | 40 (2.0) | 2 (1.4) |
| Breast | 228 (11.5) | 4 (2.9) |
| Gastrointestinal | 177 (8.9) | 11 (7.9) |
| Gynecologic | 141 (7.1) | 5 (3.6) |
| Head & Neck | 50 (2.5) | 5 (3.6) |
| Leukemia | 129 (6.5) | 21 (15.1) |
| Lymphoma | 123 (6.2) | 19 (13.7) |
| Melanoma | 29 (1.5) | 3 (2.2) |
| Prostate | 147 (7.4) | 5 (3.6) |
| Kidney | 162 (8.1) | 5 (3.6) |
| Sarcoma | 4 (0.2) | 0 (0.0) |
| Testicular | 1 (0.1) | 0 (0.0) |
| Thoracic | 114 (5.7) | 17 (12.2) |
| Thyroid | 43 (2.2) | 4 (2.9) |
| Skin (Other) | 298 (15.0) | 13 (9.4) |

**Supplemental Table 4.** Predictors of severe eGFR discrepancy. Univariable and multivariable logistic regression model for severe group defined as eGFR_CYS_ more than 50% lower than eGFR_CRE_ and eGFR_CYS_ <30 ml/min/1.73m^2^. *Chronic medication use was defined within 1 year prior to baseline; corticosteroid use was defined within 30 days of baseline. Abbreviations: eGFR_CRE-CYS_ = estimated Glomerular Filtration Rate calculated using the 2021 race-free combined serum creatinine and cystatin C equation, ACEi/ARB = Angiotensin Converting Enzyme Inhibitor/Angiotensin Receptor Blocker.

| Covariates | Severe discrepancy group (N=139)<br>Vs. Reference group (N= 1409) |  |  |  |  |  |
| --- | --- | --- | --- | --- | --- | --- |
|  | Univariable |  |  | Multivariable |  |  |
|  | OR | 95% CI | p-value | Adj OR | 95% CI | p-value |
| **Age (per 10 years)** | **1.16** | **1.02, 1.32** | **0.031** | 1.03 | 0.86, 1.25 | 0.743 |
| **Male Sex** | 0.83 | 0.58, 1.18 | 0.308 | 0.81 | 0.51, 1.29 | 0.38 |
| **White Race** | 1.35 | 0.87, 2.15 | 0.197 | 1.84 | **1.06, 3.29** | **0.033** |
| **Body mass index** |  |  |  |  |  |  |
| Normal weight | REF | — |  | — | — |  |
| Underweight | 1.52 | 0.43, 4.20 | 0.463 | 1.18 | 0.26, 4.43 | 0.82 |
| Overweight | 1.19 | 0.77, 1.89 | 0.446 | **2.22** | **1.26, 4.01** | **0.007** |
| Obese | 0.91 | 0.53, 1.57 | 0.743 | **1.98** | **0.99, 4.00** | **0.053** |
| **Smoking** | **1.50** | **1.05, 2.12** | **0.024** | 0.77 | 0.48, 1.21 | 0.26 |
| **Comorbidities** |  |  |  |  |  |  |
| Hypertension | **2.39** | **1.42, 4.31** | **0.002** | 0.62 | 0.28, 1.40 | 0.24 |
| Coronary Artery Disease | **3.24** | **2.23, 4.80** | **<0.001** | 1.14 | 0.66, 1.98 | 0.64 |
| Diabetes Mellitus | **5.36** | **3.53, 8.42** | **<0.001** | 1.71 | 0.98, 3.02 | 0.061 |
| Cirrhosis | **5.21** | **2.92, 9.02** | **<0.001** | **2.80** | **1.31, 5.98** | **0.008** |
| Human Immunodeficiency Virus | 0.98 | 0.37, 2.13 | 0.963 |  |  |  |
| Malnutrition | 1.23 | 0.74, 1.97 | 0.406 |  |  |  |
| Thyroid disease | 1.70 | 1.16, 2.45 | 0.005 | 1.17 | 0.90, 1.51 | 0.245 |
| **Medication Use\*** |  |  |  |  |  |  |
| ACEi/ARB | 0.99 | 0.70, 1.41 | 0.968 |  |  |  |
| Proton Pump Inhibitors | **3.15** | **2.04, 5.07** | **<0.001** | 1.06 | 0.58, 1.98 | 0.85 |
| Diuretics | **5.34** | **3.27, 9.28** | **<0.001** | 1.71 | 0.87, 3.50 | 0.13 |
| Corticosteroid use | **6.67** | **4.62, 9.64** | **<0.001** | **2.38** | **1.49, 3.80** | **<0.001** |
| **Baseline Labs** |  |  |  |  |  |  |
| eGFR<sub>CRE-CYS</sub> | 0.95 | 0.94, 0.96 | **<0.001** | 0.98 | 0.97, 0.99 | **<0.001** |
| Albumin (g/dL) |  |  |  |  |  |  |
| ≥4.0 | REF | — |  | — | — |  |
| 3.0-3.9 | **12.7** | **7.13, 23.7** | **<0.001** | **3.91** | **2.00, 7.96** | **<0.001** |
| <3.0 | **54.4** | **31.0, 102** | **<0.001** | **12.3** | **6.14, 26.0** | **<0.001** |
| Hemoglobin (g/dL) |  |  |  |  |  |  |
| ≥12.0 | REF | — |  | — | — |  |
| 10.0-11.9 | **3.77** | **1.76, 8.57** | **<0.001** | 1.50 | 0.64, 3.78 | 0.36 |
| ≤10.0 | **31.3** | **17.0, 64.6** | **<0.001** | **3.59** | **1.64, 8.44** | **<0.001** |

**Supplemental Table 5.** Predictors of Vancomycin level > 30μg/dL. Univariable and multivariable logistic regression model predicting Vancomycin trough level > 30 μg/dL. eGFR discrepancy defined as eGFR_CYS_ > 30% lower than eGFR_CRE_. Abbreviations: eGFR_CRE-CYS_ = estimated Glomerular Filtration Rate calculated using the 2021 race-free combined serum creatinine and cystatin C equation, ACEi/ARB = Angiotensin Converting Enzyme Inhibitor/Angiotensin Receptor Blocker.

| Covariates | Predictors of Vancomycin level > 30µg/dL(N=55)<br>Vs. <30µg/dL (N=227) |  |  |  |  |  |
| --- | --- | --- | --- | --- | --- | --- |
|  | Univariable |  |  | Multivariable |  |  |
|  | OR | 95% CI | p-value | Adj OR | 95% CI | p-value |
| **Age (per 10 years)** | **0.79** | **0.65, 0.97** | **0.023** | **0.73** | **0.57, 0.93** | **0.010** |
| **Male Sex** | 1.32 | 0.73, 2.38 | 0.36 | 1.47 | 0.64, 3.73 | 0.39 |
| **White Race** | 1.54 | 0.71, 3.72 | 0.30 | 1.38 | 0.61, 3.48 | 0.460 |
| **eGFR discrepancy** | **2.89** | **1.40, 6.58** | **0.006** | **2.30** | **1.05, 5.51** | **0.047** |
| **Body mass index** | 1.04 | 0.99, 1.09 | 0.096 | 1.04 | 0.98, 1.09 | 0.18 |
| **Smoking** | 0.91 | 0.50, 1.64 | 0.75 |  |  |  |
| Comorbidities |  |  |  |  |  |  |
| Hypertension | 1.41 | 0.60, 3.90 | 0.47 |  |  |  |
| Coronary Artery Disease | 1.27 | 0.67, 2.56 | 0.48 |  |  |  |
| Diabetes Mellitus | **3.28** | **1.36, 9.77** | **0.016** | 2.38 | 0.91, 7.47 | 0.098 |
| Cirrhosis | 0.94 | 0.36, 2.16 | 0.89 |  |  |  |
| Malnutrition | 0.49 | 0.14, 1.30 | 0.19 |  |  |  |
| Thyroid disease | 0.86 | 0.44, 1.63 | 0.66 |  |  |  |
| Medication Use* |  |  |  |  |  |  |
| ACEi/ARB | 0.96 | 0.53, 1.74 | 0.90 |  |  |  |
| Proton Pump Inhibitors | 1.27 | 0.53, 3.51 | 0.62 |  |  |  |
| Diuretics | 1.76 | 0.76, 4.83 | 0.22 |  |  |  |
| Corticosteroids | **3.51** | **1.86, 7.03** | **<0.001** | **2.91** | **1.47, 6.02** | **0.0030** |
| Baseline Labs |  |  |  |  |  |  |
| eGFR<sub>CRE-CYS</sub> | 0.99 | 0.98, 1.00 | 0.043 | 0.99 | 0.97, 1.00 | 0.032 |
| Albumin (g/dL) |  |  |  |  |  |  |
| ≥4.0 | - | - | - |  |  |  |
| 3.0 to <4.0 | 2.73 | 0.72, 17.9 | 0.20 |  |  |  |
| <3.0 | 3.05 | 0.85, 19.6 | 0.14 |  |  |  |
| Hemoglobin (g/dL) |  |  |  |  |  |  |
| ≥12.0 | - | - | - |  |  |  |
| 10.0 to <12.0 | 2.57 | 0.40, 50.4 | 0.40 |  |  |  |
| ≤10.0 | 5.05 | 1.00, 92.1 | 0.12 |  |  |  |

**Supplemental Table 6.**
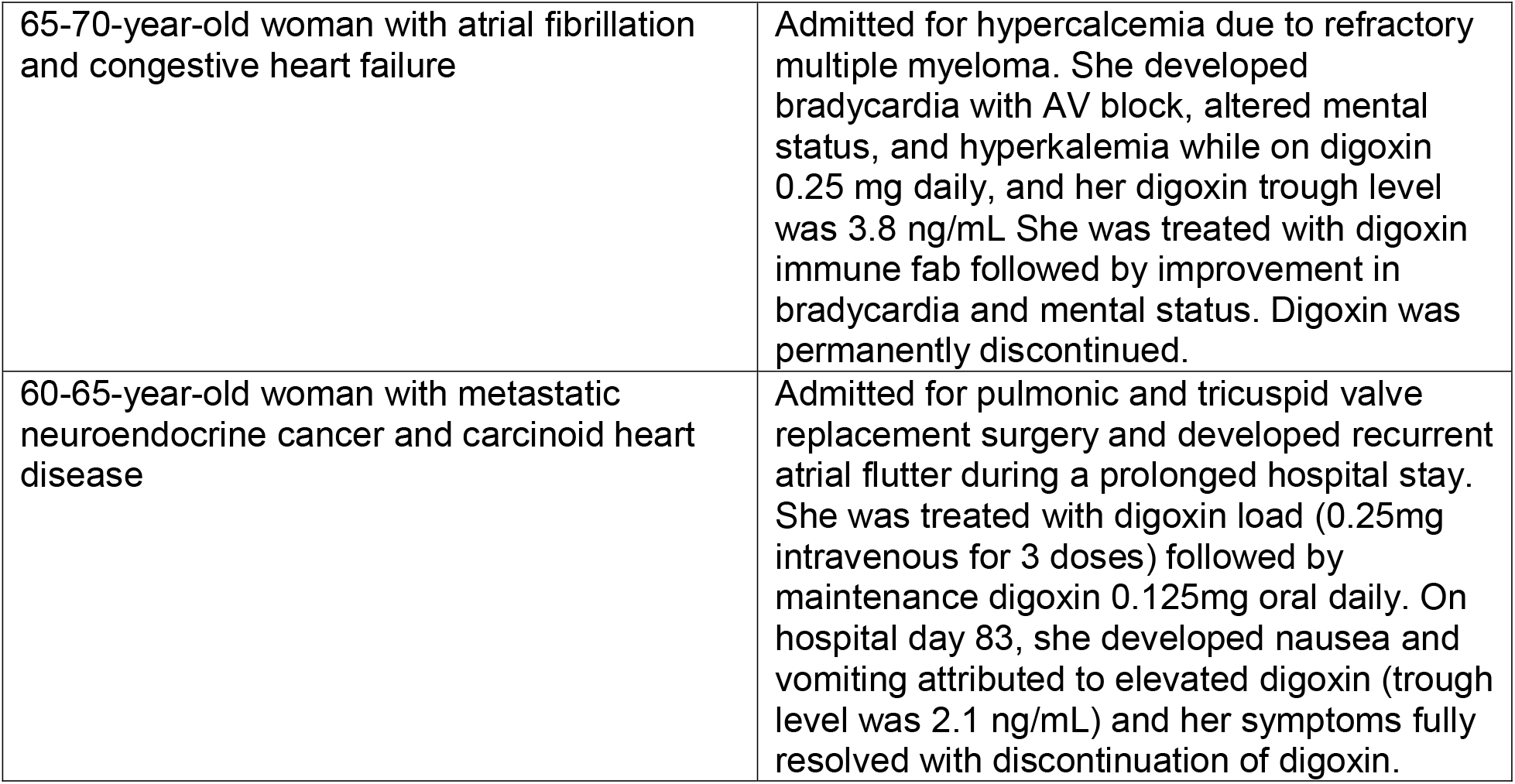
Deidentified case summaries of clinical digoxin toxicity.

**Supplemental Table 7.** Predictors of 30-day mortality. Univariable and multivariable Cox model for 30-day mortality. eGFR discrepancy defined as eGFR_CYS_ > 30% lower than eGFR_CRE_. *Chronic medication use was defined within 1 year prior to baseline; corticosteroid use was defined within 30 days of baseline. In all cases the population median was imputed for missing variables (body mass index was missing for 479 participants, serum albumin was missing for 72 participants, hemoglobin was missing for 46 participants). Abbreviations: eGFR_CRE-CYS_ = estimated Glomerular Filtration Rate calculated using the 2021 race-free combined serum creatinine and cystatin C equation, ACEi/ARB = Angiotensin Converting Enzyme Inhibitor/Angiotensin Receptor Blocker.

| Covariates | 30-day mortality |  |  |  |  |  |
| --- | --- | --- | --- | --- | --- | --- |
|  | Univariable |  |  | Multivariable |  |  |
|  | HR | 95% CI | p-value | Adj HR | 95% CI | p-value |
| **Age (per 10 years)** | 1.01 | 0.89, 1.14 | 0.93 | 1.04 | 0.90, 1.18 | 0.614 |
| **Male Sex** | 0.80 | 0.56, 1.13 | 0.20 | 1.22 | 0.85, 1.75 | 0.28 |
| **White Race** | 1.60 | 0.99, 2.57 | 0.054 | **1.71** | **1.04, 2.79** | **0.034** |
| **Body mass index** |  |  |  |  |  |  |
| Normal (18.5 - 24.9) | REF | — |  | — | — |  |
| Under weight (<18.5) | 1.20 | 0.47, 3.07 | 0.70 | 0.97 | 0.37, 2.53 | 0.96 |
| Overweight (25 - 29.9) | 0.96 | 0.64, 1.44 | 0.84 | 1.06 | 0.69, 1.62 | 0.80 |
| Obese (≥ 30) | **0.57** | **0.33, 0.98** | **0.041** | 1.15 | 0.66, 2.00 | 0.62 |
| **eGFR discrepancy** | **7.57** | **5.12, 11.2** | **<0.001** | **1.97** | **1.29, 3.01** | **0.002** |
| **Smoking** | 1.22 | 0.86, 1.71 | 0.26 |  |  |  |
| Comorbidities |  |  |  |  |  |  |
| Hypertension | 0.86 | 0.57, 1.31 | 0.49 |  |  |  |
| Coronary Artery Disease | 2.10 | 1.46, 3.03 | **<0.001** | 0.99 | 0.66, 1.51 | 0.98 |
| Diabetes Mellitus | 2.46 | 1.68, 3.60 | **<0.001** | 1.03 | 0.67, 1.59 | 0.89 |
| Cirrhosis | 2.70 | 1.62, 4.49 | **<0.001** | 1.11 | 0.64, 1.93 | 0.70 |
| Human Immunodeficiency Virus | 1.11 | 0.49, 2.53 | 0.80 |  |  |  |
| Malnutrition | 0.61 | 0.34, 1.11 | 0.11 |  |  |  |
| Thyroid disease | 1.27 | 0.87, 1.83 | 0.21 |  |  |  |
| Medication Use* |  |  |  |  |  |  |
| ACEi/ARB | **0.71** | **0.50, 1.00** | **0.049** | 0.90 | 0.60, 1.34 | 0.59 |
| Proton Pump Inhibitors | **2.18** | **1.42, 3.35** | **<0.001** | 1.20 | 0.74, 1.94 | 0.47 |
| Diuretics | **2.00** | **1.32, 3.04** | **0.001** | 0.67 | 0.40, 1.11 | 0.12 |
| Corticosteroids | **5.02** | **3.56, 7.07** | **<0.001** | **1.58** | **1.09, 2.30** | **0.017** |
| Baseline Labs |  |  |  |  |  |  |
| eGFR<sub>CRE-CYS</sub> | **0.98** | **0.97, 0.99** | **<0.001** | **0.99** | **0.98, 1.00** | **0.027** |
| Albumin (g/dL) |  |  |  |  |  |  |
| ≥4.0 | REF | — |  | — | — |  |
| 3.0 to <4.0 | **16.3** | **6.32, 42.0** | **<0.001** | **7.17** | **2.62, 19.6** | **<0.001** |
| <3.0 | **96.3** | **39.2, 237** | **<0.001** | **31.8** | **11.8, 86.2** | **<0.001** |
| Hemoglobin (g/dL) |  |  |  |  |  |  |
| ≥12.0 | REF | — |  | — | — |  |
| 10.0 to <12.0 | **6.10** | **2.45, 15.2** | **<0.001** | 2.00 | 0.77, 5.22 | 0.15 |
| ≤10.0 | **28.8** | **12.6, 65.5** | **<0.001** | **2.93** | **1.18, 7.25** | **0.020** |

